# Effect of Surgically Acquired Neurological Deficits on Mortality Among Patients with Brain Metastases

**DOI:** 10.1101/2022.11.22.22282635

**Authors:** Yilong Zheng, Sophie Jia Lin Xie, Kejia Teo, Vincent Diong Weng Nga, Tseng Tsai Yeo, Mervyn Jun Rui Lim

## Abstract

**Purpose:** To evaluate the association between surgically acquired neurological deficits and mortality among patients who underwent surgical resection of brain metastases.

**Methods:** Patients who underwent surgical resection of brain metastases at our institution between 2011 and 2019 were included. Surgically acquired neurological deficits were defined as dysarthria/aphasia, ataxia, hemiparesis, and visual field loss. A Cox proportional hazards model adjusting for potential confounders was constructed to evaluate whether surgically acquired neurological deficits were independently associated with a higher risk of overall mortality.

**Results:** A total of 153 patients were included in the analysis. 3.3% (5 patients) had a surgically acquired neurological deficit. On univariate time-to-event analysis, there was no statistically significant association between the development of a surgically acquired neurological deficit and mortality (HR=1.12; 95% CI=0.15, 8.24; p=0.910). On multivariate time-to-event analysis adjusting for potential confounders, there was also no statistically significant association between the development of a surgically acquired neurological deficit and mortality (HR=1.53; 95% CI=0.20, 11.9; p=0.683).

**Conclusion:** The development of a surgically acquired neurological deficit was not associated with overall mortality. Although this conclusion differs from other studies in the literature, the goal of surgical resection remains unchanged – to resect as much tumor as possible while still preserving neurological function.

## Introduction

Brain metastases are the most common type of brain malignancies in adults,^1^ occurring in 20% to 40% of cancer patients throughout the course of their disease.^2^ Among adults, the commonest sites of origin of cerebral metastases are the lung and breast.^3^ Surgical resection is central to the management of brain metastasis as it confers survival benefit,^4-7^ allows for the relief of symptoms,^8^ and allows for the establishment of a definitive diagnosis.^8^ However, there is also a significant risk of a surgically acquired neurological deficit.

The incidence of a surgically acquired neurological deficit has been reported to be approximately 6%.^9,10^ Preservation of neurological function is important due to its potential implications on function and quality of life.^11^ Surgically acquired neurological deficits have also recently been shown to increase the risk of mortality.^12^ Medikonda et al. reported a statistically significant association between surgically acquired neurological deficits and overall mortality among patients who underwent surgical resection of a solitary brain metastasis.^12^ However, while the findings of Medikonda et al. are significant in that they were the first to show that surgically acquired neurological deficits increase the risk of mortality among patients with solitary brain metastases, their findings have thus far not been validated. Therefore, we aimed to evaluate the association between surgically acquired neurological deficits and mortality among patients who underwent surgical resection of brain metastases.

## Methods

### Study design

This was a retrospective study of patients who underwent surgical resection of brain metastases at our institution between March 2011 and December 2019. Institutional ethics approval was obtained from the local institutional review board prior to the commencement of the study (National Healthcare Group Domain Specific Review Board; Reference Number 2020/00358), and a waiver of informed consent was granted as this study posed no more than minimal risk to participants.

### Cohort selection

The operating theatre records database was accessed to retrieve the National Registration Identity Card numbers (NRICs, which act as the national identification number), date of surgery, provisional diagnosis, operation summaries, and surgical codes of all patients who underwent surgery for a brain tumor. The medical records of the patients were then retrieved and screened for inclusion in the study. The inclusion criteria were (a) patients who had a histologically verified metastasis to the brain, and (b) patients who were 18 years old and above at the time of surgery. The exclusion criteria were (a) patients who presented only with leptomeningeal or skull metastases, and (b) patients who were followed up for less than 28 days.

### Data collection

Clinical data were collected using a standardized data collection form. Variables collected include (1) demographics (including age at the time of surgical resection, sex, and ethnicity), (2) details of the extent of disease (including the presence of systemic disease at the time of surgery and the size of the largest brain metastasis), (3) duration of hospitalization, (4) site of the primary tumor (defined as the 5 commonest sites and ‘others’), (5) and presence of preoperative and postoperative neurological deficits, and (6) whether the patient passed away. Preoperative and postoperative neurological deficits were defined as dysarthria/aphasia, ataxia, hemiparesis, or visual field loss.^12^ A surgically acquired neurological deficit was defined as a neurological deficit that developed only after surgical resection.

### Statistical analysis

The baseline characteristics of the patients were reported using mean and standard deviation for continuous variables and count numbers and percentages for categorical variables. Hypothesis testing was conducted using the Fisher’s exact test. A p-value of 0.05 or less was taken to be statistically significant.

To ascertain whether the presence of a surgically acquired neurological deficit was associated with overall mortality, Cox regression models were constructed, adjusting for age and variables that had a statistically significant association with presence of surgically acquired neurological deficits on univariate analysis. A p-value of 0.05 or less was taken to be statistically significant. For patients who demised during the follow-up period, time-to-mortality was defined as the duration between the first surgical resection and the date of death as documented in the electronic medical records. For patients who did not demise during the follow-up period, time-to-mortality was defined as the duration between the date of the first surgical resection of and the date of the latest follow-up. All data analyses were conducted using R Studio Version 1.2.5042.

## Results

### Baseline characteristics of the study population

The baseline characteristics of patients with and without surgically acquired neurological deficit(s) were reported in Table 1. A total of 153 patients met the inclusion criteria for this study. The mean (SD) age was 59.5 (11.7) years, and 81 (52.9%) patients were female. The incidence of a surgically acquired neurological deficit was 3.3% (5 patients). Details on the clinical course of the 5 patients with a surgically acquired neurological deficit were presented in Table 2.

**Table 1:** Baseline characteristics of patients with and without surgically acquired neurological deficits

| Variable | No surgically<br>acquired<br>neurological<br>deficit (n=148,<br>96.7%) | Surgically<br>acquired<br>neurological<br>deficit (n=5,<br>3.3%) | Total<br>(n=153) | p-value |
| --- | --- | --- | --- | --- |
| Age; mean (SD) | 59.4 (11.7) | 61.4 (12.4) | 59.5 (11.7) | 0.659 |
| Female; n (%) | 76 (51.4) | 5 (100.0) | 81 (52.9) | 0.061 |
| Ethnicity; n (%) |  |  |  | **0.041** |
| Chinese | 103 (69.6) | 2 (40.0) | 105 (68.6) |  |
| Malay | 24 (16.2) | 0 (0.0) | 24 (15.7) |  |
| Indian | 7 (4.7) | 0 (0.0) | 7 (4.6) |  |
| Others | 14 (9.5) | 3 (60.0) | 17 (11.1) |  |
| Systemic disease; n (%) | 95 (64.2) | 2 (40.0) | 97 (63.4) | 0.356 |
| Size of largest brain metastasis (cm); mean (SD) | 3.1 (1.1) | 3.6 (1.5) | 3.1 (1.1) | 0.251 |
| Duration of hospitalization (days); mean (SD) | 19.4 (20.3) | 58.2 (29) | 20.7 (21.6) | **0.002** |
| Site of primary tumor; n (%) |  |  |  | 0.526 |
| Lung | 59 (39.9) | 3 (60.0) | 62 (40.5) |  |
| Breast | 37 (25.0) | 1 (20.0) | 38 (24.8) |  |
| Kidney | 6 (4.1) | 0 (0.0) | 6 (3.9) |  |
| Colorectum | 17 (11.5) | 0 (0.0) | 17 (11.1) |  |
| Skin | 6 (4.1) | 1 (20.0) | 7 (4.6) |  |
| Others | 23 (15.5) | 0 (0.0) | 23 (15.0) |  |
| Preoperative neurological deficit; n (%) |  |  |  |  |
| Dysarthria/aphasia | 20 (13.5) | 2 (40.0) | 22 (14.4) | 0.151 |
| Ataxia | 18 (12.2) | 1 (20.0) | 19 (12.4) | 0.490 |
| Hemiplegia | 45 (30.4) | 2 (40.0) | 47 (30.7) | 0.643 |
| Visual field deficit | 8 (5.4) | 0 (0.0) | 8 (5.2) | 1.000 |
| Location of largest tumor; n (%) |  |  |  | 0.439 |
| Cerebellum | 30 (20.3) | 0 (0.0) | 30 (19.6) |  |
| Parietal | 28 (18.9) | 0 (0.0) | 28 (18.3) |  |
| Frontal | 49 (33.1) | 3 (60.0) | 52 (34.0) |  |
| Occipital | 11 (7.4) | 0 (0.0) | 11 (7.2) |  |
| Temporal | 15 (10.1) | 1 (20.0) | 16 (10.5) |  |
| Parieto-temporal | 1 (0.7) | 0 (0.0) | 1 (0.7) |  |
| Parieto-occipital | 7 (4.7) | 1 (20.0) | 8 (5.2) |  |
| Subcortical | 2 (1.4) | 0 (0.0) | 2 (1.3) |  |
| Fronto-parietal | 1 (0.7) | 0 (0.0) | 1 (0.7) |  |
| Temporal-occipital | 3 (2.0) | 0 (0.0) | 3 (2.0) |  |
| Sellar | 1 (0.7) | 0 (0.0) | 1 (0.7) |  |

Abbreviations: SD, standard deviation.

**Table 2:** Clinical course of patients who had surgically acquired neurological deficits

| Patient | Site of primary tumor | Location of resected brain metastasis | Immediate postoperative complications | Surgically acquired neurological deficit | Status at latest follow-up | Duration of follow-up (days) |
| --- | --- | --- | --- | --- | --- | --- |
| 1 | Lung | Temporal | Surgical site hematoma requiring evacuation | Dysarthria (resolved by POD12) | Alive | 47 |
| 2 | Lung | Parieto-occipital | Deep vein thrombosis | Hemiplegia (resolved POD5) | Demised | 66 |
| 3 | Breast | Frontal | Surgical site hematoma requiring evacuation, surgical site infection, cerebral abscess, venous sinus thrombosis, seizure | Hemiplegia (not resolved) | Alive | 83 |
| 4 | Skin | Frontal | None | Expressive aphasia (not resolved) | Alive | 97 |
| 5 | Lung | Frontal | Urinary tract infection, steroid-induced hyperglycemia | Expressive aphasia (resolved POD4) | Alive | 118 |

Abbreviations: POD, postoperative day.

### Association between surgically acquired neurological deficit and

On univariate time-to-event analysis, there was no statistically significant association between the development of a surgically acquired neurological deficit and mortality (HR=1.12; 95% CI=0.15, 8.24; p=0.910) (Table 3). On multivariate time-to-event analysis adjusting for potential confounders, there was also no statistically significant association between the development of a surgically acquired neurological deficit and mortality (HR=1.53; 95% CI=0.20, 11.9; p=0.683) (Table 3).

**Table 3:** Cox-proportional hazards analysis evaluating the association between presence of a surgically acquired neurological deficit and overall mortality

| Exposure | Crude HR (95% CI) | p-value | Adjusted HR (95% CI)<sup>1</sup> | p-value |
| --- | --- | --- | --- | --- |
| Surgically acquired neurological deficit |  |  |  |  |
| No | — |  | — |  |
| Yes | 1.12 (0.15, 8.24) | 0.910 | 1.53 (0.20, 11.9) | 0.683 |

<sup>1</sup>Adjusted for age, sex, ethnicity, and duration of hospitalization.
Abbreviations: HR, hazard ratio; CI, confidence interval.

On subgroup analysis of patients who presented with solitary brain metastasis (86 patients, 56.2%), there was no statistically significant association between surgically acquired neurological deficit and overall mortality on both univariate (HR=1.12; 95% CI=0.15, 8.26; p=0.909) and multivariate analysis (HR=1.51; 95% CI=0.19, 11.8; p=0.695).

## Discussion

In this study of patients who underwent surgical resection of brain metastases, we showed that there was no statistically significant association between the development of a surgically acquired neurological deficit and overall mortality. Our findings differ from the findings of Medikonda et al.^12^ In their study of patients who underwent surgical resection of a solitary brain metastasis, Medikonda et al.^12^ reported that a surgically acquired neurological deficit was significantly associated with a higher risk of overall mortality. In our study, we followed Medikonda et al.’s definition of neurological deficit,^12^ except that we considered dysarthria a neurological deficit as well.

The association between a surgically acquired neurological deficit and mortality has been postulated to be due to the confounding effect of the quality of life.^13^ Patients with surgically acquired neurological deficits have a higher risk of mortality because these patients also tend to have poorer quality of life, and a poorer quality of life has been associated with a higher risk of mortality.^13^ However, the retrospective nature of Medikonda et al.’s^12^ and our study make the assessment of the quality of life difficult, and therefore it is unclear whether the association between surgically acquired neurological deficits may be attributed to the confounding effect of quality of life. Prospective studies that assess and control for the quality of life in the analysis may help clarify this association.

However, one should be cognizant of the goal of surgical treatment when interpreting studies on the effect of surgically acquired neurological deficits on mortality. As Liau pointed out, whether surgically acquired neurological deficits actually impact survival is practically irrelevant, as the goal of surgical resection has been and should continue to be to resect as much tumor as possible while still maintaining neurological function, regardless of whether or not the length of overall survival is significantly impacted by the acquisition of a new neurological deficit.^14^

There are several limitations to our study. First, the small sample size of our cohort limited the statistical power of our analyses, with only 5 patients having a surgically acquired neurological deficit. Also, as our study included only patients from a single institution, the results may not be generalizable to other institutions. Lastly, we do not rule out the possibility of undocumented surgically acquired neurological deficits, and therefore the incidence of surgically acquired neurological deficits may be lower than the actual incidence reported in this study.

## Conclusions

The development of a surgically acquired neurological deficit was not associated with overall mortality. Although this conclusion differs from other studies in the literature, the goal of surgical resection remains unchanged – to resect as much tumor as possible while still preserving neurological function.

## Data Availability

All data produced in the present study are available upon reasonable request to the authors

## Ethical Approval

Institutional ethics approval was obtained from the institutional review board prior to study initiation (National Healthcare Group Domain Specific Review Board; Reference Number 2020/00358).

## Informed Consent

A waiver of informed consent was granted for this study as this study.

